# APOB to estimated APOB ratio for screening for the APOE2 genotype

**DOI:** 10.64898/2026.01.29.26345063

**Authors:** C Auger, M Sampson, R Zubiran, J Cole, A Wolska, J Otvos, A Sniderman, AT Remaley

## Abstract

**Background:** Familial dysbetalipoproteinemia (FDB) is a genetic lipoprotein disorder that can develop in patients homozygous for the APOE2 genotype (ε2/ε2). It is associated with decreased clearance of remnant lipoproteins and increased atherosclerotic cardiovascular disease (ASCVD) risk disproportionate to their level of LDL-C. A goal of this study was to develop a screening test for the ε2/ε2 genotype based on routinely available lipid tests and to determine those at most risk for ASCVD.

**Methods:** After assembly of a primary prevention cohort from the UK Biobank (n= 269,895), gene array and exome data was utilized to classify patients as being ε2/ε2 genotype positive or negative. Lipid profiles and APOB levels were extracted and the number of ASCVD events was tabulated during a 15-year follow-up period.

**Results:** Using a newly developed equation for estimating APOB (eAPOB) with lipid panel test results, the ratio of measured APOB to eAPOB was better than any other individual lipid test or ratio for identifying patients with the ε2/ε2 genotype (AUC: APOB/eAPOB: 0.990 (0.986-0.994), nonHDL-C/APOB: 0.961 (0.952-0.970), APOB: 0.955 (0.949-0.961), VLDL/TG: 0.788 (0.771-0.804)). The majority of ε2/ε2 patients could be identified with the APOB/eAPOB ratio even before they expressed the FDB phenotype with elevated TG and nonHDL-C. The PCE or PREVENT risk equations were the most accurate method for identifying higher risk patients (AUC: PREVENT: 0.690 (0.637-0.742), PCE: 0.697 (0.645-0.749)).

**Conclusion:** The APOB/eAPOB ratio can be used to accurately identify the ε2/ε2 genotype and conventional risk equations are the best method for determining those at risk for ASCVD.

## Introduction

Familial dysbetalipoproteinemia (FDB) is a genetic disorder that results in the plasma accumulation of remnant lipoproteins, which are partially lipolyzed chylomicrons and Very Low-Density Lipoproteins (VLDL) (1–3). It is associated with increased risk of atherosclerotic cardiovascular disease (ASCVD) and can occur in individuals who are homozygous for ε2 gene allele, which encodes for the apolipoproteinE2 isoform. Less often, it occurs in patients with rare mutations in the *APOE* gene or in other lipoprotein-related genes (1–4). Apolipoprotein E (APOE) is an exchangeable-type apolipoprotein that serves as a ligand for various receptors and is found on VLDL and other lipoproteins (5). Compared to the more common APOE3 isoform, APOE2 has decreased affinity for proteoglycans (6), Low-Density Lipoprotein (LDL) Receptor and LDL Receptor Related Protein 1 (7), which together mediate the removal of remnant lipoproteins from the circulation (8). Due to delayed conversion of VLDL to LDL and other compensatory changes, most individuals with the ε2/ε2 genotype show low levels of LDL-Cholesterol (LDL-C) and Apolipoprotein B (APOB) (1–3).

Historically, the FDB phenotype was diagnosed with lipoprotein electrophoresis by detecting β-VLDL, an abnormal cholesterol-enriched remnant lipoprotein, which migrates slower than normal VLDL (9). Later, a ratio of VLDL-C/TG >0.3 (mg/dL) as determined by ultracentrifugation, along with some other lipid parameters, was used to biochemically define FDB (9). Recently, an equation based on the lipid panel test results and APOB has been described for estimating VLDL-C without the need for ultracentrifugation (10), but the VLDL-C/TG ratio is not specific for FDB. More than half of all patients with a VLDL-C/TG >0.3, as determined by ultracentrifugation, are not homozygous for APOE2 and have lipoprotein remnant accumulation from other causes besides FDB (2). A wide variety of lipid tests and lipid ratios have been used to identify for the FDB phenotype, but the nonHDL-C/APOB ratio is usually the most effective (1, 11, 12).

Only a minority of patients (<20%) with the ε2/ε2 genotype go on to develop severe dyslipidemia and are therefore categorized as having the FDB phenotype and are at increased risk for ASCVD. The presence of a “second hit”, such as obesity, insulin resistance or hypothyroidism, has been suggested to be necessary for the clinical phenotype to appear (13–17). Other known FDB triggers are hypothyroidism, alcohol use, renal disease and exogenous estrogen use (15–17).

Because past efforts have mostly focused on identifying FDB patients after they have already developed a significant dyslipidemia, the goal of this study was to determine if the ε2/ε2 genotype could possibly be identified earlier. Identifying these patients before they develop the FDB phenotype and dyslipidemia could improve clinical management by advising them in the prevention of obesity and diabetes, and or in the avoidance of other known triggers for FDB. In addition, we also wanted to investigate how to best determine ASCVD risk in individuals who are known to be ε2/ε2 genotype positive, regardless of their FDB phenotype status.

## Methods

Permission to use test results and ASCVD outcome data from the UK Biobank was obtained under application #97500. Patient consent and IRB review was performed by the original UK Biobank study, which received approval from the NorthWest Multi-Centre Research Ethics Committee (11/NW/0382) (18). Use of deidentified test results in this study was considered nonhuman subject research (45 CFR 46.104) and exempt from IRB review.

Incident ASCVD events over a 15-year follow-up period was defined to include the UK Biobank algorithmically defined outcomes for myocardial infarction and stroke, as well as the following ICD 10 groups related to ASCVD: I20(0,1,8,9), I21(0,1,2,4,9), I22(0,1,2,8,9), I23(0-8), I24(0,1,8,9), I25(0-9), I60(0-9), I61(0-6, 8,9), I62(0,1,9), I63(0-6,8,9), I64(0), I65(0,1,2,8,9), I66(0-3,8,9), I70(0,2,8,9), I73(8,9), E10(5), E11(5), E14(5). Patients on lipid-lowering medication, those with an ASCVD event at baseline, and those without available genomic or lipid data were excluded from analysis. The remaining cohort of 298,249 subjects were randomly split into a training dataset for model derivation (n=149,307) and a validation dataset (n=148,942).

PLINK (v1.90b6.21) was utilized to extract two SNPs, rs7412 and rs29358, from Affymetrix UK BiLEVE Axiom data array and whole exome sequencing. Concordant genotypes from both data types were retained and discordant results removed (<1%). If genotype results were only available from one source, results were still retained. Individuals were classified as ε2/ε2, if they were homozygous for the thymine (T) allele at rs7412 and showed no variant at rs29358.

Standard lipid panel (total cholesterol, high density lipoprotein cholesterol [HDL-C], Triglycerides [TG]) and APOB were measured directly as previously described (18). LDL-C and VLDL-C were calculated by the enhanced Sampson (eS) equation that includes APOB as an independent variable with the version of the equation based on normolipidemic individuals (19). The PCE and PREVENT risk (10-year) scores were calculated as previously described (20–22). The mean PCE/PREVENT scores for the ε2/ε2 population, split by gender, was determined.

Expected ASCVD events was calculated based on the mean risk scores multiplied by 1.5. This was compared to the actual event rate in the ε2/ε2 population. To compare to the non-ε2/ε2 population, 100 PCE/PREVENT matched non-ε2/ε2 control samples were identified for each ε2/ε2 patient above and below the Deming regression line for the plot of Non-HDLC and TG. An observed ASCVD event rate was then calculated for these controls and compared by Chi-square analysis to predicted event rate for the PCE/PREVENT scores and to the observed event rate in the ε2/ε2 population.

Unless otherwise indicated, all results are expressed as mean values ±1 SD. Group comparisons were performed using Student’s t-test or Chi-square analysis as appropriate. Least Square regression analysis was used to fit the new equation for estimating APOB (eAPOB) from the ε3/ε3 genotype. Statistical analysis was done with JMP software. Confidence intervals for AUCs were determined utilizing the pROC package in R (23). An Excel spreadsheet for doing the various calculations used in this study, including the calculation for eAPOB, can be downloaded at the following website: (figshare.com/articles/software/Sampson_estimated_apoB/30287155?file=58519852)

## Results

The frequency of the ε2/ε2 genotype, as well as the frequency of several common dyslipidemias, was determined in a large primary prevention cohort assembled from the UK Biobank (Figure 1A). Individual patient lipid results for the log of TG and nonHDL-C were plotted and color coded to correspond to a lipoprotein phenotype classification system previously described based on these two lipid parameters (24). The dotted diagonal line is the Deming regression line which generally reflects where the greatest population density occurs for bivariate normally distributed data. In Fig. 1B, the plot is overlayed with ε2/ε2 patients that did (black circles) or did not (grey circles) develop ASCVD within the 15-year observation period.

**Figure 1:**
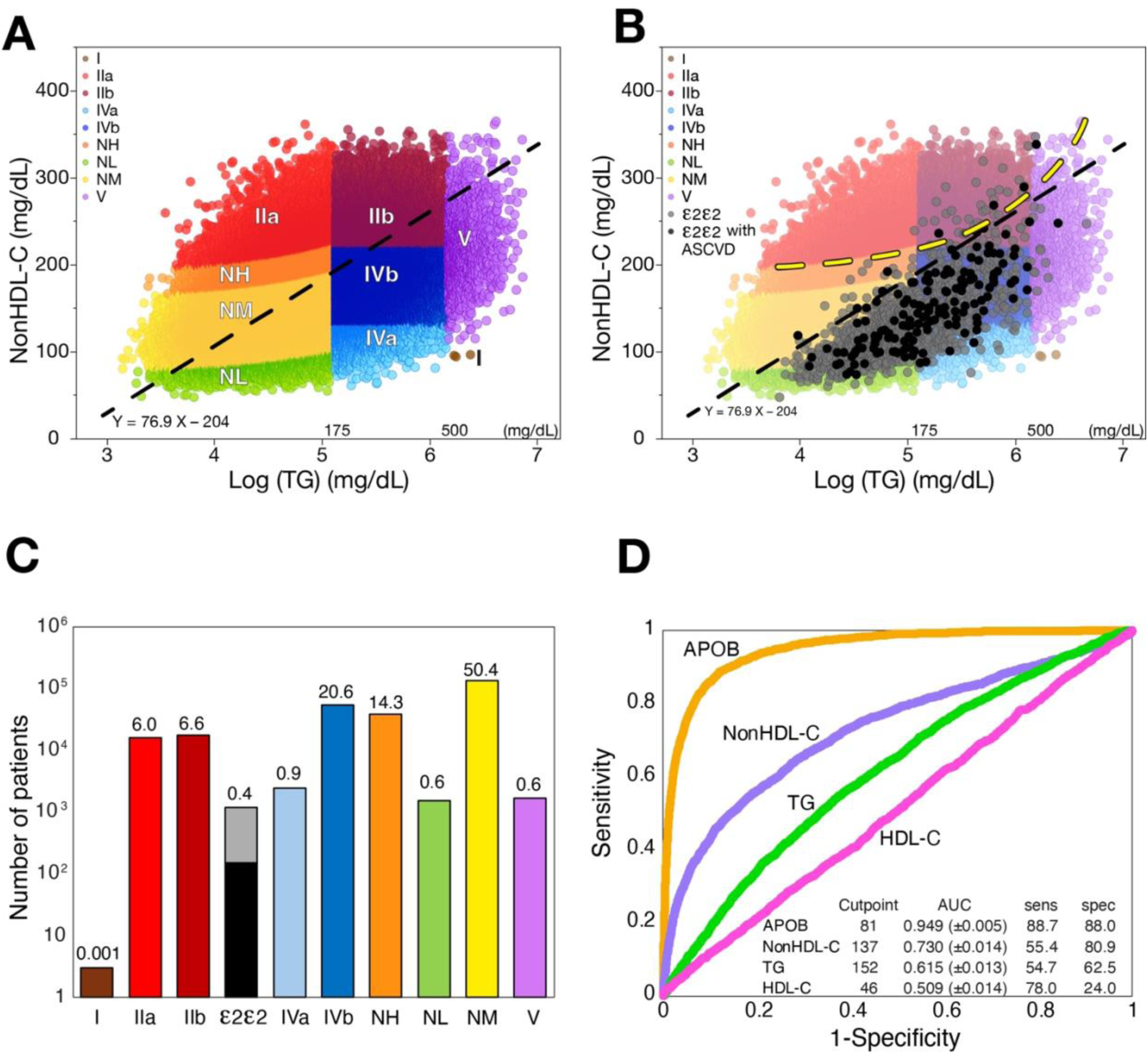
Lipoprotein distribution and lipoprotein phenotypes in UK Biobank primary prevention cohort. **(A)** Log(TG) and nonHDL-C is plotted for patients within the UKBiobank (n= 298,249) Individual points each representing a single patient are color coded by lipoprotein phenotype (Modified Fredrickson classification) (A). Diagonal dashed line is the Deming regression line. (B) Results from ε2/ε2 subjects are overlaid and those shown in black had an ASCVD event where grey points represent ε2/ε2 subjects that did not have an ASCVD event. Yellow dashed line represents LDL-C = 190 mg/dL. (C) The log of the number of each lipoprotein phenotype is plotted on the y-axis. The percent of the total population is shown above each column.

Most ε2/ε2 patients have TG levels below 500 mg/dL, with nonHDL-C values that fall below the diagonal regression line. This is because LDL-C, which is low in these patients, typically accounts for the greatest fraction of cholesterol in nonHDL-C. The region of the plot above the yellow dotted line indicates those patients with an LDL-C >190 mg/dL. Rarely did ε2/ε2 subjects have an elevated LDL-C >190 mg/dL (0.5%) or a nonHDL-C >220 mg/dL (6.2%), an alternative marker of hypercholesterolemia (21). The lipid values for the ε2/ε2 group most often overlap with non-ε2/ε2 individuals who have moderate hypertriglyceridemia (Type IV) or normolipidemia. The absolute number and percent of the population for each lipoprotein phenotype are shown in a log scale in Fig. 1C. Only about 0.4% of the population were found to have the ε2/ε2 genotype, and 13.6% of them had an ASCVD event during the follow-up period (Table 1). In contrast, hypertriglyceridemia (Type IV) and hypercholesterolemia (NH: LDL-C between 160-190 mg/dL) were both relatively common and together accounted for about a third of the population.

**Table 1.**
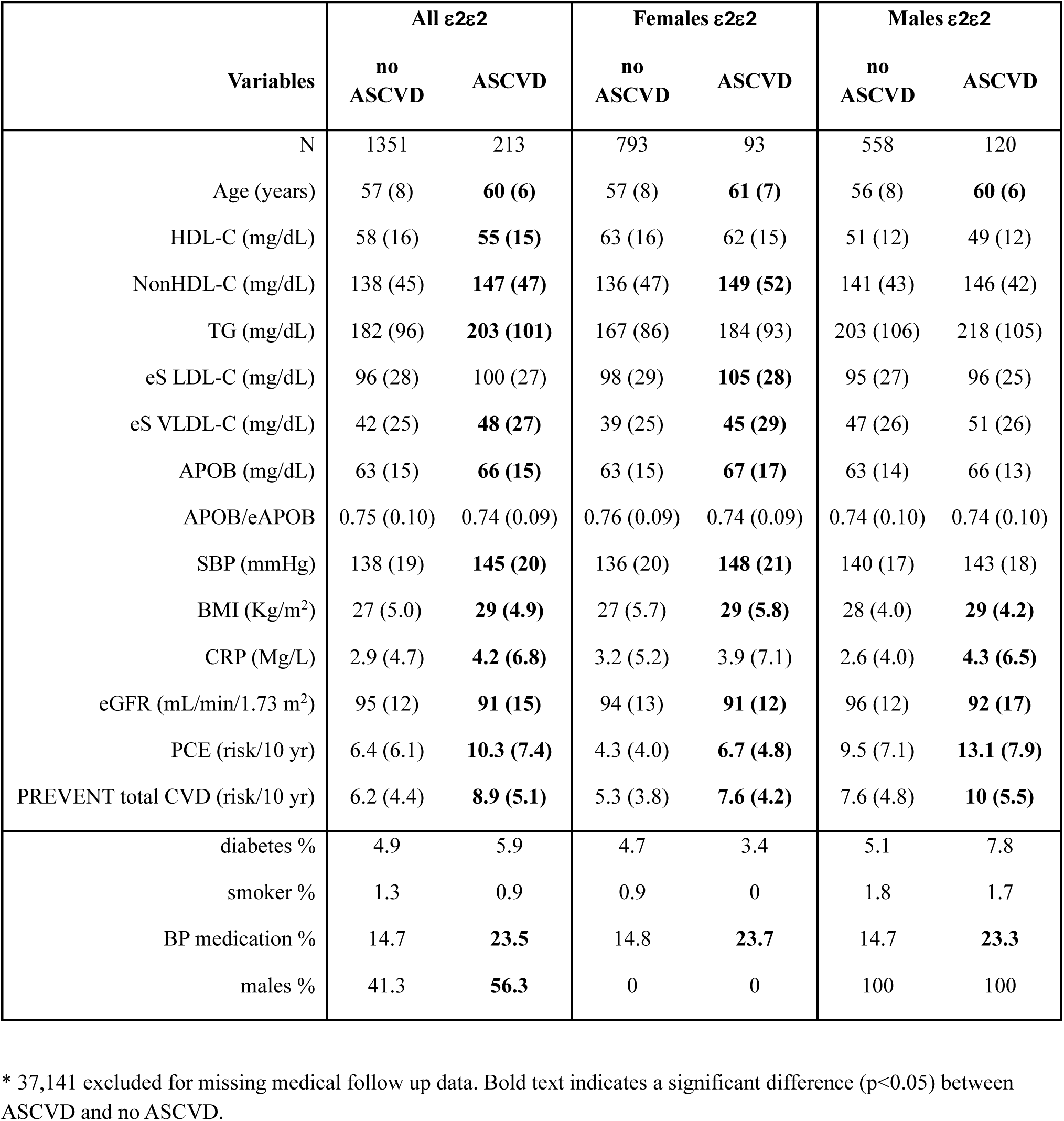

In Figure 1D, we performed ROC analysis for APOB and the tests in the standard lipid panel for identifying the ε2/ε2 genotype. APOB showed the best discrimination, and its mean value was considerably lower in subjects with the ε2/ε2 genotype (ε2/ε2: 63.5±14.8 mg/dL, non-ε2/ε2: 107.2±22.9 mg/dL, p<0.01).

The ε2/ε2 genotype is split into grey (no ASCVD events during follow-up) and black (ASCVD positive). (D) ROC curves for differentiating the ε2/ε2 genotype from non-ε2/ε2 population are shown for the indicated lipid tests. The optimum cutpoint based on Youden’s index is in inset table along with sensitivity and specificity and AUC with 95% CI shown in parentheses.

To further improve its discrimination, we developed a new equation by least square regression analysis for estimating APOB (eAPOB) to be used in conjunction with measured APOB (Fig. 2A). The following equation for eAPOB was based only on the standard lipid panel test results and derived from e3/e3 individuals in the training dataset (n=86,678).:

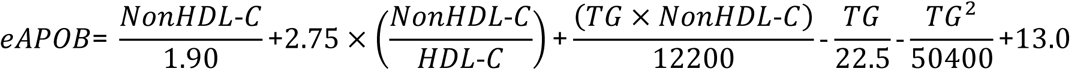

**Figure 2:**
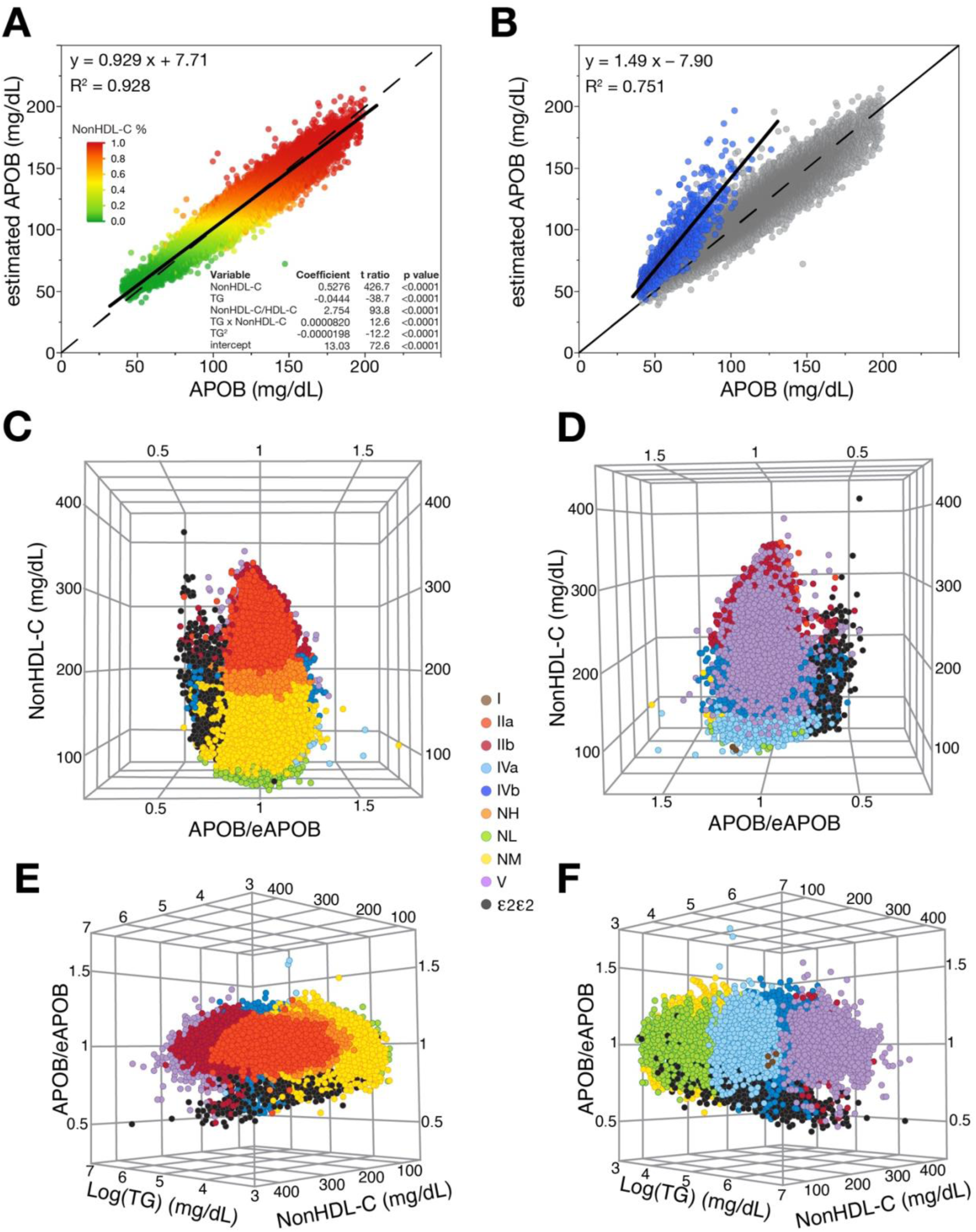
Equation for eAPOB and APOB/eAPOB ratio by lipoprotein phenotypes. (A) Correlation of eAPOB to APOB in validation dataset. Solid line represents the least squares regression line and the dashed line represents the line of identity. Results are colored by the percentile of nonHDL-C. Table inset shows regression parameters from training dataset. (B) Correlation of eAPOB to APOB in validation dataset showing non-ε2/ε2 subjects in validation data (n=148,046; grey) and ε2/ε2 individuals (n=896; blue). Solid line is the least squares regression line for ε2/ε2 individuals and dashed line is line of identity. (C-F) 3D-plots of APOB/eAPOB, non-HDL-C, and Log(TG) of the validation dataset. Color coding corresponds to lipoprotein phenotypes with same color scheme as in Figure 1.

As one can see in the inset table for the individual variables in the eAPOB equation (Fig. 2A), nonHDL-C shows the strongest association with APOB, but TG and HDL-C also contribute to its estimation. When this equation was applied to the ε2/ε2 genotype, eAPOB values were consistently higher than measured APOB (Fig. 2B). This was expected because β-VLDL carries more cholesterol than does normal VLDL, but like normal VLDL it still only has only one copy of APOB, leading to its overestimation. Next, we used eAPOB along with measured APOB to make a ratio. Conceptually, it is like the nonHDL-C/APOB ratio, in that it provides a cholesterol per APOB-particle correction based on nonHDL-C, but it is also affected by TG and HDL-C. The APOB/eAPOB ratio has a smaller population coefficient of variation than APOB alone (APOB: 23% (ε2/ε2) and 21% (non-ε2/ε2); APOB/eAPOB: 13% (ε2/ε2) and 5.7% (non-ε2/ε2)), which should help improve its discrimination ability. As can be seen in the 3D-plots of nonHDL-C, log (TG) and APOB/eAPOB (Fig. 2C-F), subjects with the ε2/ε2 genotype have a lower APOB/eAPOB ratio. Most of the ε2/ε2 genotype subjects, even those without an elevated TG or nonHDL-C, clustered in a different region of the plot compared to the other lipoprotein phenotypes.

By ROC analysis (Fig. 3A), the APOB/eAPOB ratio provided a more accurate discrimination of the ε2/ε2 genotype (AUC: 0.990 (0.986-0.994)) than APOB (AUC: 0.955 (0.949-0.961)) or the other lipid tests. It was also superior to the nonHDL-C/APOB ratio, which is used for screening for FDB (11, 12). This is most apparent in their respective positive predictive values (PPV). The PPV of APOB/eAPOB was 17.2% versus 7.6% for the nonHDL-C/APOB ratio at their optimum cut-points. We also examined a more complex prediction model using logistic regression, which included the APOB/eAPOB ratio, along with APOB and eAPOB, but it was only slightly better than just the ratio. Because of its low prevalence, the new APOB/eAPOB ratio still had a relatively low PPV despite its good AUC. We, therefore, examined several other potential cut-points. Its PPV could be improved to 50% with a negative predictive value (NPV) of 99.9% by changing the cut-point for positivity to <0.849, which lowers its sensitivity from 95.6% to 87.8%. The eSVLDL-C/TG ratio, which was designed to detect all causes of dysbetalipoproteinemia (10), had the lowest overall specificity and sensitivity for detecting the ε2/ε2 genotype. We also tested the various lipid tests and ratios by sex but did not detect any major sex specific differences for identifying the ε2/ε2 genotype.

**Figure 3.**
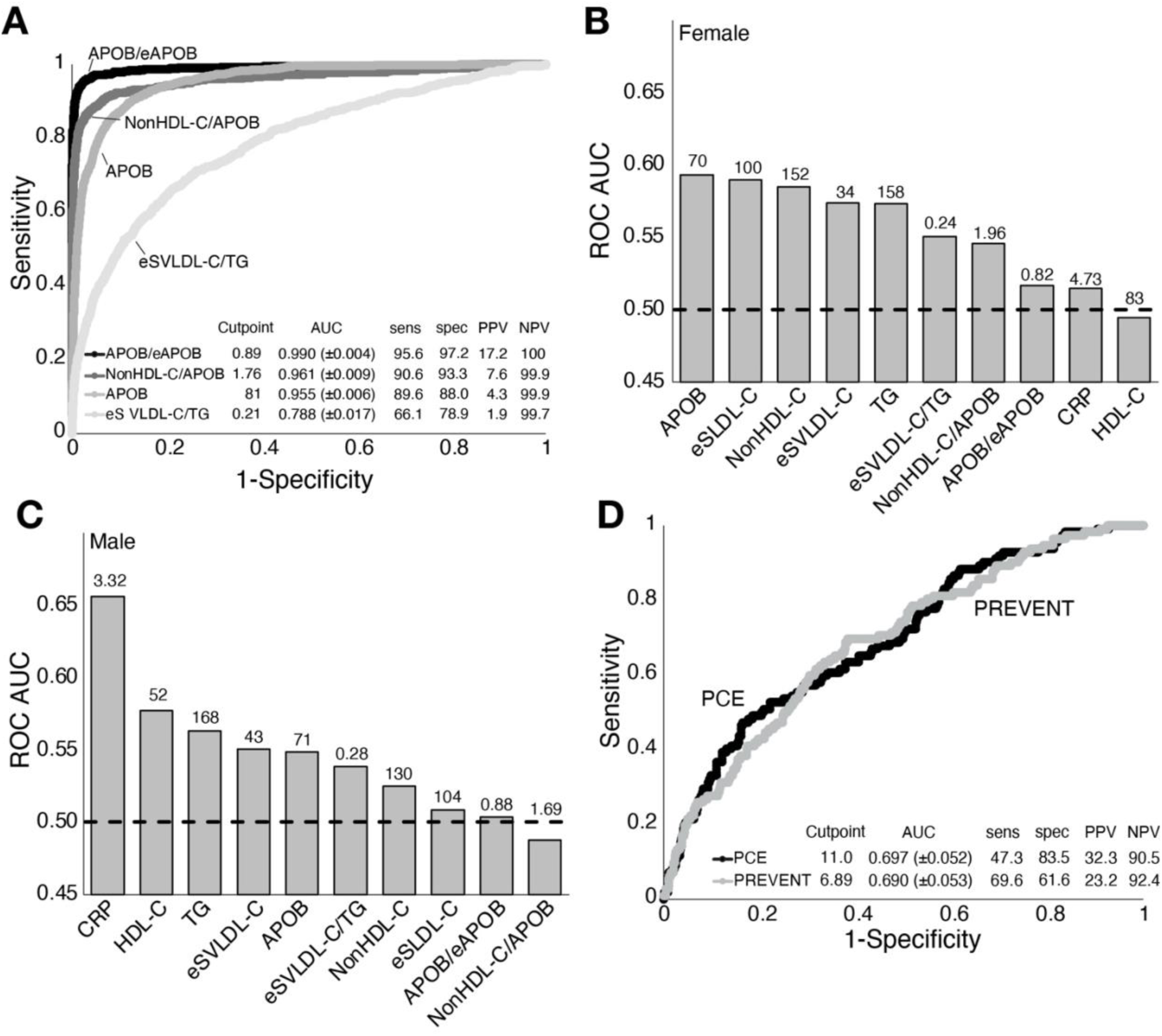
ROC analysis of lipid tests and ratios for the identification of ε2/ε2 genotype and their risk for ASCVD. (A) ROC curve analysis from validation set (n=148,942) for detection of ε2/ε2 genotype by indicated lipid test or ratio. Table inset shows AUC (± 95% confidence interval), sensitivity, specificity, NPV, and PPV for each test. (B) AUC scores for indicated test for females for predicting ASCVD. Numbers above bars indicate optimum cutpoint. (C) AUC scores for indicated test for males for predicting ASCVD. Numbers above bars indicate optimum cutpoint. (D) ROC analysis of PCE and PREVENT for predicting ASCVD. Table inset indicates optimum cutpoints, AUC (± 95% confidence interval), sensitivity, specificity, NPV, and PPV.

Next, the different lipid tests and ratios were examined by sex for predicting ASCVD events in patients who were positive for the ε2/ε2 genotype (Fig. 3B, 3C). All the individual tests showed relatively poor performance and their rank order varied by sex. APOB (AUC: 0.59) was the best in females, whereas hsCRP (AUC: 0.66) was the best in males. Both the APOB/eAPOB and nonHDL-C/APOB ratios were ranked relatively low in their effectiveness as ASCVD risk markers for both sexes. Besides lipids, other types of risk factors including age (Table 1) varied significantly between ε2/ε2 genotype patients with and without ASCVD. We, therefore, investigated PCE (Fig. 3D), the current US recommended risk score that includes age, sex and different lipid and non-lipid risk factors (21), for calculating a 10-year risk score. We also examined the newer PREVENT risk score, which besides traditional risk factors also includes eGFR (20). Both risk scores had considerably higher AUCs (PCE AUC: 0.697 (0.645-0.749); PREVENT AUC: 0.690 (0.637-0.742)) than any individual lipid risk factor or ratio, but they did not significantly differ from each other. We also examined a modified PREVENT risk score that also includes HbA1C and urine albumin/creatinine ratio, but inclusion of these additional tests did not substantially improve ASCVD prediction over the simple PREVENT risk score.

In Figure 4, we compared by sex the expected number of ASCVD events, calculated based on the mean PCE or PREVENT score of the ε2/ε2 population, to the observed ASCVD event rate that occurred in the ε2/ε2 population over the 15-year follow-up period. For females, PCE significantly underestimated the ASCVD event rate by approximately 40% ((p<0.00001), Fig. 4A); however, it did not significantly underestimate the ASCVD event rate in males (p=0.066). In contrast, the PREVENT risk score significantly underestimated the ASCVD event rate in both females (p=0.016) and males (p<0.00001).

**Figure 4.**
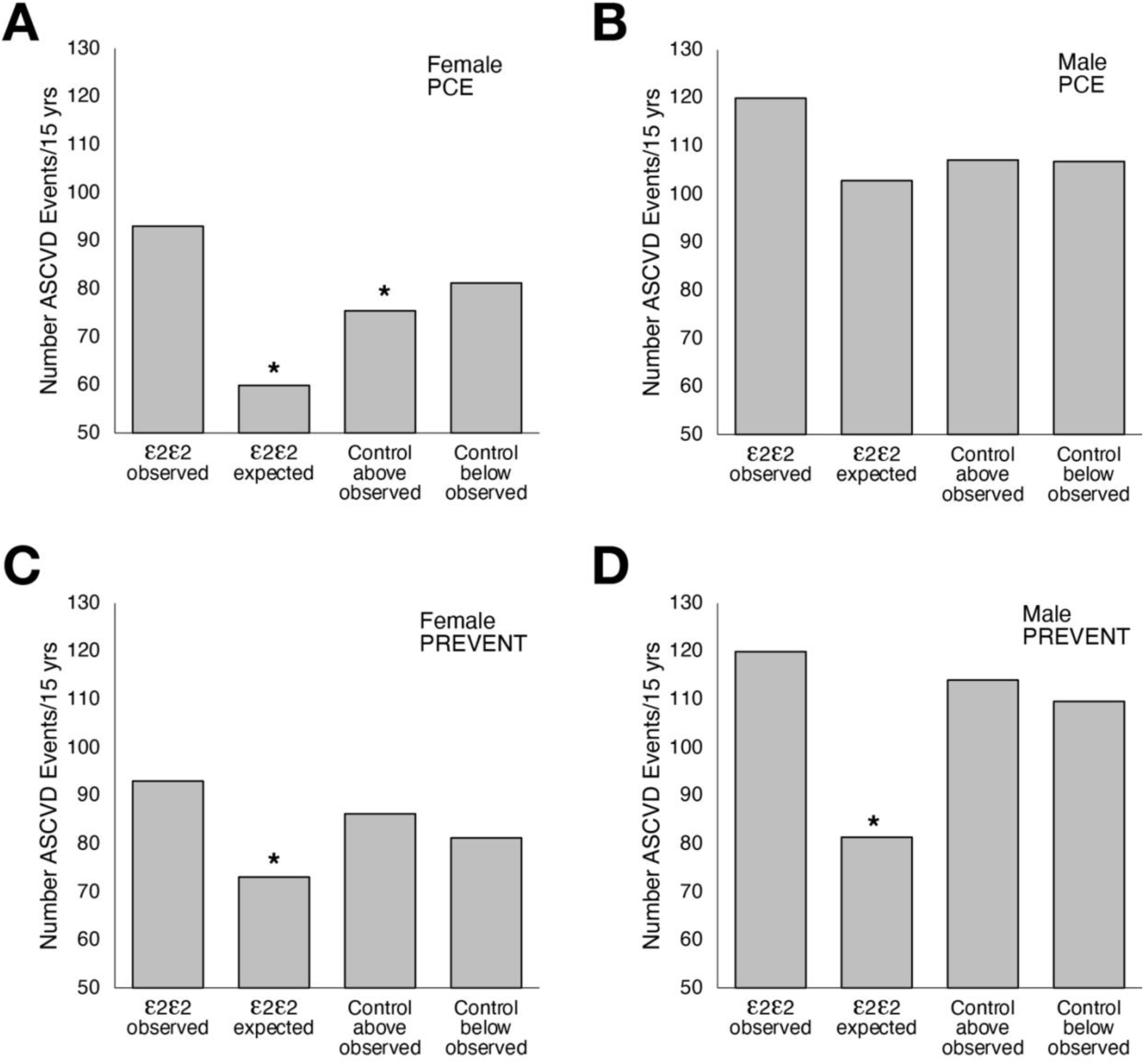
ASCVD event rate by ε2/ε2 genotype and non-2/ε2 genotype control groups. Samples with missing medical follow-up were excluded (n=37,141) and the ε2/ε2 patients (n=1,564) were divided into female (n=886, panels A and C) and male (n=678, panels B and D). “ε2/ε2 observed” indicates the ASCVD event rate for the ε2/ε2 genotype. “ε2/ε2 expected” indicates the expected ASCVD event rate in ε2/ε2 genotype group based on either their PCE (panels A and B) or PREVENT (panels C and D) risk scores. “Control (above and below) observed” indicates the observed ASCVD events during the follow-up period for the two different control groups that were designed to match the PCE and PREVENT scores in ε2/ε2 genotype group. Asterisks above bars indicates a significant (p<0.05) difference in ASCVD event rate between the “ε2/ε2 observed” group and one of the three other groups.

To determine if the underestimation of ASCVD events by the two risk scores was specific to the ε2/ε2 genotype or a function of the PCE/PREVENT scores, we made two non-ε2/ε2 genotype control groups, matched to either have the same PCE or PREVENT risk score as the ε2/ε2 genotype group (100 matches per ε2/ε2 patient; sampled for a confidence interval). In the “Control below” group, subjects were selected from below the diagonal regression line in Fig. 1B. This was done to develop a control group with approximately the same level of nonHDL-C and TG as in the ε2/ε2 genotype (Table 2). For the “Control above” group, subjects from above the regression line with relatively higher nonHDL-C but similar TG levels were selected. Despite differences in the level of the various risk factors in the two different control groups (Table 2), they had a similar number of observed ASCVD events, which was expected given their matched ASCVD risk scores. The number of observed ASCVD events in the two control groups above and below the diagonal line also did not significantly vary from the number of observed ASCVD events in the ε2/ε2 genotype group, except for a small decrease for females in the PREVENT “Control above” group. This indicates that the underestimation of ASCVD events by these risk equations is not specific to the ε2/ε2 genotype. It is instead most likely due to the miscalibration of the risk score equations for the subset of patients selected in this analysis.

**Table 2.**
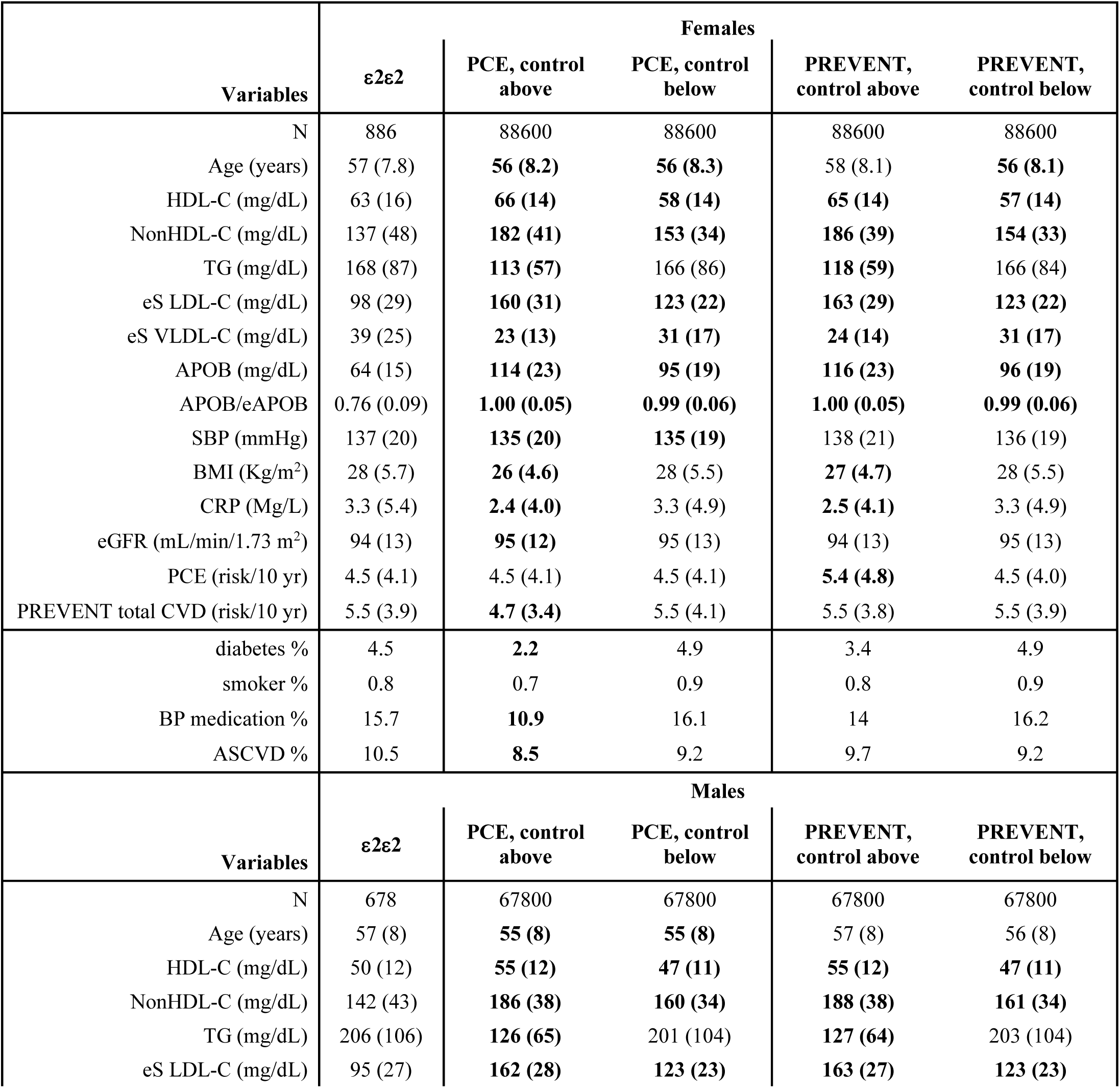
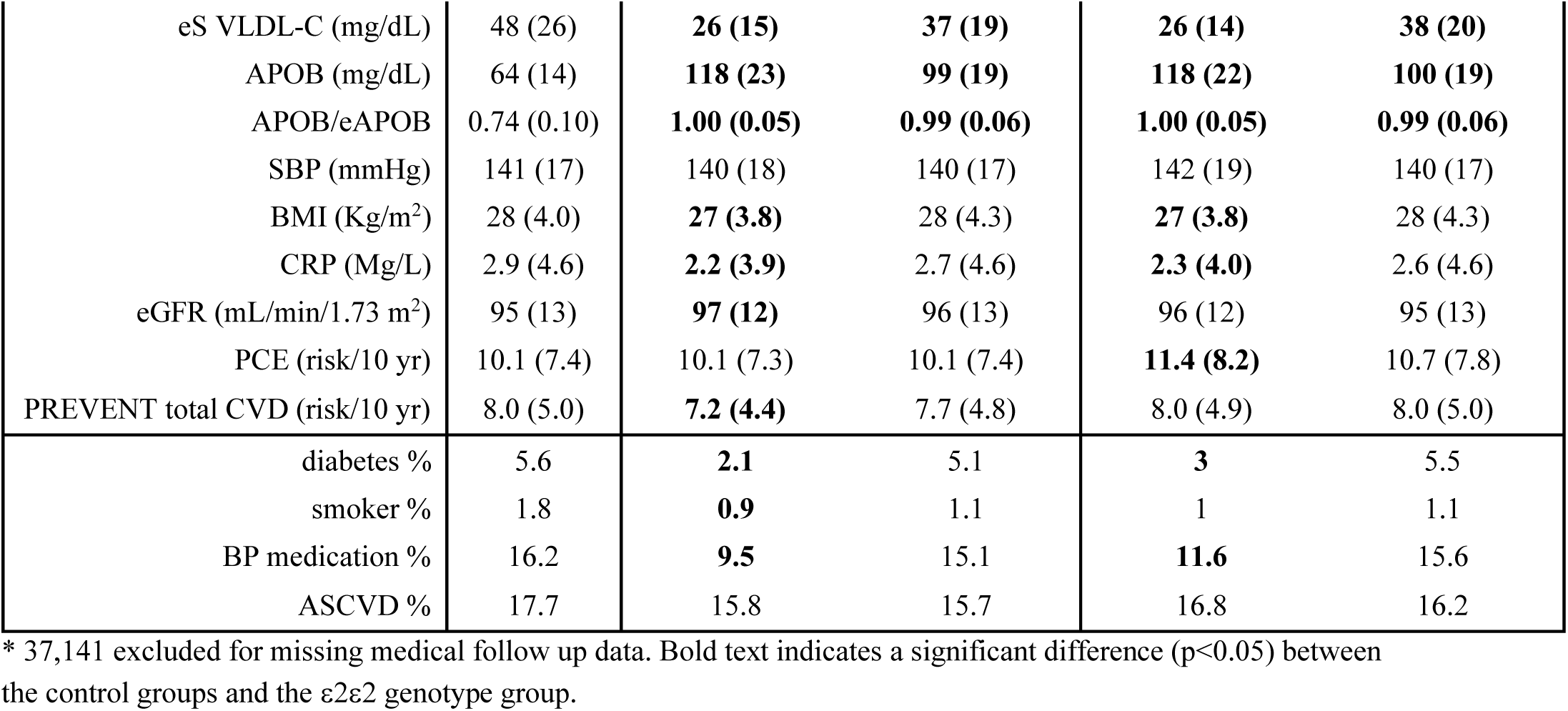

| Variables | $\epsilon 2/\epsilon 2$ | Females | | | |
| --- | --- | --- | --- | --- | --- |
|  |  | PCE, control above | PCE, control below | PREVENT, control above | PREVENT, control below |
| N | 886 | 88600 | 88600 | 88600 | 88600 |
| Age (years) | 57 (7.8) | **56 (8.2)** | **56 (8.3)** | 58 (8.1) | **56 (8.1)** |
| HDL-C (mg/dL) | 63 (16) | **66 (14)** | **58 (14)** | **65 (14)** | **57 (14)** |
| NonHDL-C (mg/dL) | 137 (48) | **182 (41)** | **153 (34)** | **186 (39)** | **154 (33)** |
| TG (mg/dL) | 168 (87) | **113 (57)** | 166 (86) | **118 (59)** | 166 (84) |
| eS LDL-C (mg/dL) | 98 (29) | **160 (31)** | **123 (22)** | **163 (29)** | **123 (22)** |
| eS VLDL-C (mg/dL) | 39 (25) | **23 (13)** | **31 (17)** | **24 (14)** | **31 (17)** |
| APOB (mg/dL) | 64 (15) | **114 (23)** | **95 (19)** | **116 (23)** | **96 (19)** |
| APOB/eAPOB | 0.76 (0.09) | **1.00 (0.05)** | **0.99 (0.06)** | **1.00 (0.05)** | **0.99 (0.06)** |
| SBP (mmHg) | 137 (20) | **135 (20)** | **135 (19)** | 138 (21) | 136 (19) |
| BMI (Kg/m<sup>2</sup>) | 28 (5.7) | **26 (4.6)** | 28 (5.5) | **27 (4.7)** | 28 (5.5) |
| CRP (Mg/L) | 3.3 (5.4) | **2.4 (4.0)** | 3.3 (4.9) | **2.5 (4.1)** | 3.3 (4.9) |
| eGFR (mL/min/1.73 m<sup>2</sup>) | 94 (13) | **95 (12)** | 95 (13) | 94 (13) | 95 (13) |
| PCE (risk/10 yr) | 4.5 (4.1) | 4.5 (4.1) | 4.5 (4.1) | **5.4 (4.8)** | 4.5 (4.0) |
| PREVENT total CVD (risk/10 yr) | 5.5 (3.9) | **4.7 (3.4)** | 5.5 (4.1) | 5.5 (3.8) | 5.5 (3.9) |
| diabetes % | 4.5 | **2.2** | 4.9 | 3.4 | 4.9 |
| smoker % | 0.8 | 0.7 | 0.9 | 0.8 | 0.9 |
| BP medication % | 15.7 | **10.9** | 16.1 | 14 | 16.2 |
| ASCVD % | 10.5 | **8.5** | 9.2 | 9.7 | 9.2 |
| Variables | $\epsilon 2/\epsilon 2$ | Males | | | |
|  |  | PCE, control above | PCE, control below | PREVENT, control above | PREVENT, control below |
| N | 678 | 67800 | 67800 | 67800 | 67800 |
| Age (years) | 57 (8) | **55 (8)** | **55 (8)** | 57 (8) | 56 (8) |
| HDL-C (mg/dL) | 50 (12) | **55 (12)** | **47 (11)** | **55 (12)** | **47 (11)** |
| NonHDL-C (mg/dL) | 142 (43) | **186 (38)** | **160 (34)** | **188 (38)** | **161 (34)** |
| TG (mg/dL) | 206 (106) | **126 (65)** | 201 (104) | **127 (64)** | 203 (104) |
| eS LDL-C (mg/dL) | 95 (27) | **162 (28)** | **123 (23)** | **163 (27)** | **123 (23)** |
| eS VLDL-C (mg/dL) | 48 (26) | **26 (15)** | **37 (19)** | **26 (14)** | **38 (20)** |
| APOB (mg/dL) | 64 (14) | **118 (23)** | **99 (19)** | **118 (22)** | **100 (19)** |
| APOB/eAPOB | 0.74 (0.10) | **1.00 (0.05)** | **0.99 (0.06)** | **1.00 (0.05)** | **0.99 (0.06)** |
| SBP (mmHg) | 141 (17) | 140 (18) | 140 (17) | 142 (19) | 140 (17) |
| BMI (Kg/m<sup>2</sup>) | 28 (4.0) | **27 (3.8)** | 28 (4.3) | **27 (3.8)** | 28 (4.3) |
| CRP (Mg/L) | 2.9 (4.6) | **2.2 (3.9)** | 2.7 (4.6) | **2.3 (4.0)** | 2.6 (4.6) |
| eGFR (mL/min/1.73 m<sup>2</sup>) | 95 (13) | **97 (12)** | 96 (13) | 96 (12) | 95 (13) |
| PCE (risk/10 yr) | 10.1 (7.4) | 10.1 (7.3) | 10.1 (7.4) | **11.4 (8.2)** | 10.7 (7.8) |
| PREVENT total CVD (risk/10 yr) | 8.0 (5.0) | **7.2 (4.4)** | 7.7 (4.8) | 8.0 (4.9) | 8.0 (5.0) |
| diabetes % | 5.6 | **2.1** | 5.1 | **3** | 5.5 |
| smoker % | 1.8 | **0.9** | 1.1 | 1 | 1.1 |
| BP medication % | 16.2 | **9.5** | 15.1 | **11.6** | 15.6 |
| ASCVD % | 17.7 | 15.8 | 15.7 | 16.8 | 16.2 |

\* 37,141 excluded for missing medical follow up data. Bold text indicates a significant difference (p<0.05) between the control groups and the $\epsilon 2\epsilon 2$ genotype group.

## Discussion

Several important findings on how to identify and clinically manage patients with the ε2/ε2 genotype were revealed from this study. First, we describe a new ratio, APOB/eAPOB, that was superior to nonHDL-C/APOB for screening for the ε2/ε2 genotype. We also found that without APOB it is not possible to accurately identify the ε2/ε2 genotype, as was previously described for the screening tests used for FDB identification (10). The new APOB/eAPOB ratio, however, not only depends upon APOB, but also on the effect of nonHDL-C, HDL-C and TG in detecting the abnormal lipid composition of APOB-containing lipoproteins in ε2/ε2 subjects.

This accounts for its improved performance over the nonHDL-C/APOB ratio, which only utilizes two lipid parameters. The APOB/eAPOB ratio is also a better diagnostic marker than just APOB, because it has a smaller population coefficient of variation while still maintaining a relatively large mean difference between the ε2/ε2 and non-ε2/ε2 genotype groups. This suggests that making a ratio between a measured risk factor and an estimated risk factor could be a general strategy for improving diagnostic test performance, if the main predictive biomarker of interest has other closely correlated laboratory tests that can be used in its estimation.

Another important finding from this study is that most ε2/ε2 genotype subjects have an abnormal lipid composition in their APOB-containing lipoproteins even before they express the FDB phenotype with an elevated TG and NonHDL-C. The current concept of a “second hit” in acutely transforming individuals with the ε2/ε2 genotype into the FDB phenotype and abruptly increasing their ASCVD risk by triggering a major qualitative and quantitative change in their lipoprotein profile may need to be reconsidered. Previous screening efforts have largely tried to identify high-risk FDB patients once they developed elevated triglycerides or some other sort of lipid abnormality. Our analysis shows that none of the individual lipid tests or ratios were accurate in identifying patients with the ε2/ε2 genotype who went on to develop an ASCVD event. Part of the explanation for the poor diagnostic performance of the lipid tests may be due to the relatively high biological variability, particularly TG, which is perhaps even more variable in individuals with the ε2/ε2 genotype like it has already been shown for FDB (25). Our analysis also indicates that, like in the general population, it is best to use a comprehensive risk score that includes age, lipids along with other risk factors for determining ASCVD risk in ε2/ε2 genotype patients despite the abnormal lipid composition of their APOB-containing lipoproteins.

Based on our analysis, individuals with the ε2/ε2 genotype, in general, do not appear to have a markedly increased risk of ASCVD when compared to other patients with similar lipids. This was evident when the observed ASCVD event rate in patients with ε2/ε2 genotype was similar to the observed ASCVD event rate in the two different non-ε2/ε2 genotype control groups, who were matched by either the PCE or PREVENT score. Because of the high ASCVD incidence in FDB (26–28), this finding is perhaps unexpected, but what is true for FDB may not necessarily apply to patients with just the ε2/ε2 genotype. In fact, large epidemiological studies that have previously shown that, in general, the ε2/ε2 genotype has an overall lower ASCVD hazard ratio than other APOE genotypes (29). Because it is genetically determined, individuals with the ε2/ε2 genotype from an early age have lower LDL-C and APOB levels, two major drivers of atherosclerosis. Thus, the early favorable lipoprotein profile in young ε2/ε2 genotype subjects may counterbalance the pro-atherogenic lipid profile that may occur later in adulthood if they start to develop the FDB phenotype.

We also observed that two commonly used ASCVD risk equations, namely PCE and PREVENT, can sometimes underestimate ASCVD risk in the ε2/ε2 genotype, which could negatively impact on the decision to use lipid-lowering therapy. Additional studies should be performed to confirm our findings, but this underestimation of risk, at least for the PREVENT equation, did not appear to be specific for ε2/ε2 genotype. It is likely due to a limitation in the risk score equations for a subset of patients used in this study with lipid profiles and risk scores like patients with ε2/ε2 genotype.

It is important to note that the screening for the ε2/ε2 genotype followed up by confirmatory genotyping could affect clinical management in other ways besides the decision on whether to use a lipid-lowering therapy. Most importantly, APOB or nonHDL-C should be the primary treatment target rather than LDL-C in these patients, because they both capture risk from other atherogenic lipoproteins besides LDL, like remnant lipoproteins, which are known to be elevated in FDB (30, 31). As can be seen in our analysis, a much larger fraction of cholesterol is also present in remnant lipoproteins (VLDL-C) in the ε2/ε2 genotype than in other subjects.

Patients identified as having the ε2/ε2 genotype could potentially benefit from lifestyle recommendations related to the prevention of obesity, diabetes, use of alcohol, and other known predisposing factors that can trigger the development of the FDB phenotype. Identifying someone with the ε2/ε2 genotype could also alter the choice of lipid-lowering therapy. Compared to other APOE genotypes, individuals with the ε2/ε2 genotype are known to show a much greater TG-lowering response when placed on a fish oil supplement (32). It may also be good to avoid the use of bile acid sequestrant in these patients, because it has been found in FDB to worsen hypertriglyceridemia (3). Finally, although the e2/e2 genotype appears to be protective against Alzheimer’s disease (33), it increases the risk for Age Related Macular Degeneration (AMD) (34). If individuals with the e2/e2 genotype develop early signs or symptoms of AMD, a vitamin supplement specifically designed for AMD should be considered given its proven benefit for eye health in the general population (35).

In summary, the new APOB/eAPOB ratio was shown to be the best screening test for identifying the ε2/ε2 genotype. Its use can potentially alter medical care by improving ASCVD risk assessment and clinical management. Our study, however, has several limitations. Although we used a large primary prevention cohort, it had limited racial diversity and thus our findings should be tested in other cohorts. We also did not include in our analysis the impact of rare APOE gene mutations, which can have a more profound effect on lipoprotein metabolism (3).

Finally, additional studies are needed to determine how to best integrate, in routine clinical practice, the APOB/eAPOB screening method in conjunction with APOE genotyping for the diagnosis and management of patients.

## Data Availability

Equations produced in the present work are contained in the manuscript. Data for this manuscript belongs to the UKBiobank.

https://figshare.com/articles/software/Sampson_estimated_apoB/30287155?file=61064338

## Acknowledgements

This research has been conducted using the UK Biobank Resource under Application 97500.

## Abbreviations

FDB: Familial dysbetalipoproteinemia
APOE: Apolipoprotein E
ASCVD: Atherosclerotic cardiovascular disease
LDL-C: Low-Density Lipoproteins-Cholesterol
APOB: Apolipoprotein B
eAPOB: Estimated APOB
nonHDL-C: NonHigh-Density Lipoprotein-Cholesterol
VLDL: Very Low-Density Lipoprotein
TG: Triglycerides
FU: Follow-up
PPV: Positive Predictive Value
NPV: Negative Predictive Value
eSVLDL-C: Very Low-Density Lipoprotein-C estimated by the Sampson equation
Genes: APOE: (Apolipoprotein E)

